# Sexual Behaviours and Sexual Health Among Middle-Aged and Older Adults in Britain: A Latent Class Analysis of The Natsal-3

**DOI:** 10.1101/2021.11.11.21265960

**Authors:** Junead Khan, Emily Greaves, Clare Tanton, Hannah Kuper, Thomas Shakespeare, Eneyi E. Kpokiri, Yun Wang, Jason J. Ong, Suzanne Day, Stephen W. Pan, Weiming Tang, Bingyi Wang, Xin Peng, Bowen Liang, Huachun Zou, Joseph D. Tucker, Dan Wu

## Abstract

**Objectives:** Population-representative studies of the sexual health of middle-aged and older adults are lacking in aging societies. This study aimed to identify latent patterns of sexual behaviours and health of people aged 45-74.

**Methods:** We conducted a latent class analysis of the National Attitudes and Sexual Lifestyles Survey (Natsal-3), a nationally representative survey conducted in Britain in 2011.

**Results:** Of the 5260 respondents aged 45-74, 49% of men and 45% of women belonged to the *Content Caseys* class who reported good sexual health. The *Infrequent Indigos* (31% of men, 44% of women) were characterized by a lack of sexual activity, reported some dissatisfaction, and were more likely to have a disability. The *Low-Functioning Lees* (12% of men, 8% of women) reported some more disability and had issues with sexual functioning and higher levels of distress. The *Multiple-Partnered Morgans* (9% of men, 2% of women) were characterized by a greater number of sexual partners and several risk behaviours.

**Conclusions:** The use of these four classes can aid in improved targeting of tailored sexual health services to improve sexual function, sexual satisfaction, reduce distress and risky behaviours among middle-aged and older adults. These services should be inclusive of the disabled community.

**Bullet points:**

- About half of these respondents were classified as *Content Caseys* who reported few issues concerning their sex lives.
- Over a third who reported no recent sexual activity, the vast majority of whom are single, were classified as *Infrequent Indigos*.
- Nearly 10% were classified *Low-Functioning Lees* due to the greater probability of reporting sexual function issues and distress.
- The rest were classified as *Multiple Partner Morgans* as a result of a high likelihood of having multiple partners and unsafe sex.

## INTRODUCTION

Longer life spans and elevated quality of care for older people have contributed to a fundamental transition about what it means to be an older adult, altering sexual biographies and behaviours.^1 2^ Although sexual activity generally decreases with age,^3^ the English Longitudinal Study of Ageing (ELSA) showed that a decent portion of middle-aged and older men and women remain sexually active.^4^ However, there is a persistent misperception that “age is a condom” and older adults aged 65 years or above do not have sex or sexual health needs. ^5^ Additionally, research on sexual health among middle-aged adults has been limited ^6^ and many sexual health studies have either explicitly excluded older participants,^7 8^ or aggregate entire subsets of middle-aged and older adults.^9 10^ There are few population-representative studies of sexual health that include these groups, even though this information is important for planning messaging campaigns, behavioural interventions, clinical programs, and health systems.^11^ There is therefore a large evidence gap on the sexual health needs of middle-aged and older adults, which themselves are heterogeneous across age groups and experience a range of sexual health needs.^11^

Previous studies have reported high levels of sexual dysfunction and dissatisfaction among older adults, where commonly reported conditions concern erectile problems for men, arousal for women, and a general decline of interest in sex.^4 12^ In addition, sexually transmitted infections (STIs) are increasing among the older age groups^13^ and few older adults report speaking to their doctor about sex at all.^12^ Thus the limited evidence we do have on older adults suggests a need to better identify and respond to older adults’ needs for sexual health services in ways that address their unique, age-related concerns. Characterizing these concerns and identifying the range of need for this diverse demographic will improve the ability of care providers and policymakers to prioritize interventions.

We sought to evaluate the sexual behaviours and health outcomes of middle-aged and older adults and identify sexual health needs among this group. Using the National Survey of Sexual Attitudes and Lifestyles (Natsal-3), a rare example of a population representative sexual health study that included older adults, we conducted a latent class analysis (LCA) to create and examine identifiable classes. LCAs have recently gained attention through their ability to characterize differing levels of risk^14^ and health outcomes, including in sexual health.^15^ Past LCAs that cluster symptoms and biomarkers to study disease profiles offer a model for translating demographic and behavioural characteristics into clinical guides that could push forward a personalized approach to sexual health service provision.^16^ The purpose of this LCA sub-analysis was to characterize the sexual health of this population and parse out differing sets of sexual health needs that can be used to develop and tailor appropriate services and interventions.

## METHODS

### Overview

This study is a secondary analysis of data from Britain’s third National Survey of Sexual Attitudes and Lifestyles (Natsal-3), a nationally representative probability sample survey conducted in 2010-12.^17^ We included middle-age people (i.e., people who are aged 45-64, a period of human adulthood that immediately precedes the onset of old age) and older adults who are aged 65 or above. We broadened our scope to include middle-aged adults because the 40s are associated with a transition from early to older adulthood in British life and culture and important biological changes (e.g., female and male menopause). Other studies focusing on ageing have frequently included participants in their 40s and 50s.^18 19^ We used LCAs to identify unobserved (i.e., latent) classes of middle-aged and older people with respect to their sexual health characteristics using sexual health items.^20^ This method uses responses to survey questions in order to identify new subgroups that are defined by observed variables. We then compared socio-demographic, health-related, and help-seeking variables across classes. In doing this we hope to capture a better understanding of the population structure of middle-aged and older adults, the diversity of their experiences and needs, and to inform potential strategies for intervention.

### Participants and procedures

Natsal-3 was the third cross-sectional, roughly decennial nationwide population representative household survey focused on sexual lifestyle and attitudes in Britain. The survey recruited 15,162 men and women age 16 to 74 in 2010-2012. Natsal-3 is one of the first of its kind to survey older adults with this breadth and comprehensiveness and the next survey (Natsal-4), which is planned to be fielded for spring 2022, will extend only up to 59 years of age.^21^ Methods describing the design and administration of Natsal-3 have been published previously.^17^ The technical report and questionnaire are available online (www.natsal.ac.uk). The survey used a combination of face-to-face computer-assisted personal interview and computer-assisted self-interview for the more sensitive questions, including questions about sexual health and sexual behaviours. Verbal consent was obtained from each participant before they commenced the survey.

### Survey instruments and definitions

The survey asked participants for socio-demographic information. Participants were also asked about their opinions towards their general health status, treatment history for depression in the past 12 months, sexual behaviours and practices, sexual function, feeling of sexual satisfaction, sexual distress, and history of STIs. For the purpose of this analysis, we have defined “unsafe sex” as the incidence of unprotected sex with two or more partners. “Low sexual function” was derived from a five-point Likert scale developed and validated by the Natsal-3 study team.^22^ Additionally, participants were also asked to identify whether they had a limiting disability, defined as any long-standing illness, disability or infirmity that limits their activities in any way. The phrasing of these questions can be found in both the Natsal-3 study^17^ and Appendix Table I.

### Statistical analysis

Analyses used complex survey functions in order to incorporate the weighting, clustering and stratification of the data. Weights were derived to account for unequal probabilities of selection and non-response and corrected for differences in gender, age and regional distribution according to the UK 2011 census for this analysis.^17^ We firstly reported differences in socio-demographic characteristics, sexual behaviours and outcomes for the entire age range of the sample using Chi-squared tests. We then focused our comparison on the 45-54, 55-64, and 65-74 age groups. Analyses were carried out using JMP® Pro, Version 15.0 (SAS Institute Inc., Cary, NC).

### Latent class analyses

We hypothesized that men and women may show varying patterns of sexual practices and outcomes, and we stratified the latent class analyses by sex. We included a set of variables that were key measurements of sexual behaviour (i.e., frequency of sex in last four weeks, number of partners in past year), higher-risk sexual behaviours (unsafe sex in past year, purchasing sex in past year), and sexual health outcomes (STI diagnosis in the past five years, sexual function, sexual satisfaction, and sexual distress). Each latent class model item was coded as a binary variable to improve interpretability of model results. Identification of variables for inclusion was an iterative process, and the final list of variables was determined based on discussions among the authors and published literature.

We determined the optimal number of classes based on Akaike Information Criteria (AIC), Bayesian Information Criteria (BIC), and the negative log-likelihood. Lower values indicate better fit in all cases, though BIC has been found to be the most reliable metric.^23^ Interpretability and rationale for the combination of variables for classes were also considered. In naming and characterizing the classes, a probability greater than 50% for a certain item was considered an indication that members of a given class were more likely to report that risk factor. After identifying latent classes, we cross-tabulated on socio-demographic, behavioural and lifestyle variables by classes to investigate the individuals who make up the classes for potential tailoring of interventions.

### Ethical approval

The Natsal-3 study was approved by the Oxfordshire Research Ethics Committee A (reference: 09/H0604/27).

## RESULTS

### Demographic Backgrounds

Data from 9902 respondents were eligible for this sub-analysis. Among them, 5260 respondents age 45-74 were the focus of this study, of which 2233 were men and 3027 were women. Table 1 presents demographic characteristics of participants alongside data for those age <45 years. Among those aged 45-74 years, most identified as white (92% for both men and women) and over half (55% of men, 51% of women) were married or in a civil partnership. In addition, 28% of men and 30% of women had a limiting disability.

**Table 1.** Demographic characteristics of male and female participants in NATSAL-3, by age group, 2012, UK, N = 15162 (Created by the authors)

| Class Label | Men (%) |  |  |  | Women (%) |  |  |  |
| --- | --- | --- | --- | --- | --- | --- | --- | --- |
|  | < 45<br>(n =<br>4060) | 45-54<br>(n =<br>794) | 55 - 64<br>(n =<br>772) | 65 - 74<br>(n =<br>667) | < 45<br>(n =<br>5842) | 45-54<br>(n =<br>1123) | 55 - 64<br>(n =<br>1030) | 65 - 74<br>(n =<br>874) |
| Ethnicity |  |  |  |  |  |  |  |  |
| White | 85 | 91 | 95 | 96 | 85 | 89 | 94 | 96 |
| Mixed | 3 | 1.0 | 0.5 | 0.6 | 3 | 2 | 0.9 | 0.3 |
| Asian | 7 | 4 | 2 | 2 | 6 | 3 | 3 | 1 |
| Black | 4 | 3 | 1 | 1 | 4 | 5 | 1 | 1 |
| Other † | 1.6 | 0.8 | 0.5 | 0.0 | 1.6 | 0.9 | 1 | 0.2 |
| Sexual Identity |  |  |  |  |  |  |  |  |
| Heterosexual/Straight | 97 | 96 | 97 | 99 | 96 | 97 | 99 | 99 |
| Gay/lesbian | 2 | 2 | 1 | 0.2 | 1 | 1 | 0.8 | 0.1 |
| Bisexual | 1 | 1 | 1 | 0.5 | 2 | 1 | 0.0 | 0.3 |
| Other | 0.4 | 0.7 | 0.4 | 0.3 | 0.3 | 0.7 | 0.2 | 0.2 |
| Relationship Status |  |  |  |  |  |  |  |  |
| Married or civil partnership | 23 | 50 | 54 | 62 | 28 | 49 | 55 | 50 |
| Living with a partner | 18 | 12 | 7 | 3 | 20 | 9 | 5 | 3 |
| In a steady relationship, not cohabiting | 19 | 11 | 9 | 4 | 20 | 13 | 5 | 2 |
| No steady relationship | 39 | 24 | 27 | 26 | 31 | 27 | 33 | 41 |
| Academic Qualifications |  |  |  |  |  |  |  |  |
| No academic qualifications | 10 | 17 | 37 | 52 | 8 | 17 | 38 | 35 |
| Academic qualifications typically<br>gained at age 16 years $\bar{x}$ | 32 | 40 | 25 | 18 | 32 | 40 | 33 | 27 |
| Studying for or have attained<br>further academic qualifications | 52 | 42 | 37 | 29 | 54 | 41 | 27 | 18 |
| Socio-Economic Class ‡ | | | | | | | | |
| Manager/professional | 32 | 44 | 41 | 26 | 31 | 39 | 31 | 15 |
| Intermediate | 34 | 30 | 31 | 29 | 27 | 25 | 29 | 20 |
| Semi-routine/routine | 34 | 30 | 31 | 29 | 27 | 25 | 29 | 20 |
| No job currently | 3 | 4 | 9 | 27 | 8 | 11 | 18 | 50 |
| Student | 16 | 1 | 0.0 | 0.4 | 16 | 0.9 | 0.3 | 0.2 |
| Respondent has a longstanding illness, disability or infirmity |  |  |  |  |  |  |  |  |
| None | 81 | 62 | 45 | 42 | 78 | 60 | 49 | 41 |
| Non-Limiting | 9 | 16 | 24 | 25 | 11 | 13 | 20 | 26 |
| Limiting | 10 | 22 | 31 | 32 | 11 | 27 | 31 | 33 |
| Opinion of own health |  |  |  |  |  |  |  |  |
| Very Good | 46 | 35 | 26 | 24 | 46 | 36 | 32 | 25 |
| Good | 43 | 45 | 42 | 41 | 42 | 42 | 39 | 41 |
| Fair | 10 | 16 | 22 | 28 | 10 | 16 | 20 | 24 |
| Bad | 1 | 4 | 7 | 7 | 2 | 5 | 7 | 7 |
| Very Bad | 0.4 | 0.8 | 3 | 1 | 0.3 | 1 | 2 | 2 |
| Quintile of Index of Multiple Deprivation |  |  |  |  |  |  |  |  |
| 1 (least deprived) | 18 | 23 | 22 | 26 | 17 | 23 | 23 | 25 |
| 2 | 19 | 22 | 25 | 26 | 19 | 21 | 23 | 25 |
| 3 | 19 | 19 | 21 | 19 | 20 | 20 | 20 | 18 |
| 4 | 23 | 18 | 16 | 17 | 22 | 17 | 20 | 16 |
| 5 (most deprived) | 21 | 19 | 17 | 12 | 22 | 20 | 14 | 17 |

All participants. \*Participants aged > 16. ¨English General Certificate of Secondary Education or equivalent. ‡National Statistics Socio-Economic
Classification. † Other includes Chinese

### Behavioural Characteristics

Appendix Table II presents behavioural characteristics of participants aged >45. Of these respondents, fewer men reported no sexual partners in the past year (27% of men, 37% of women) and more men reported having 2 or more partners in the past year (10 % of men, 3% of women). Men also reported more often engaging in all other sexual practices, with the exception of masturbation in the past month which was more commonly reported by women (50% of men, 60% women). Men were more commonly engaged in risky sexual behaviours such as unsafe sex in the past year (5% of men, 2% of women). Similar trends were observed for those diagnosed with an STI in the last five years (1% of men, 0.6% of women) while 17% of men and 16% of women reported a low sexual function score. All p values for this table were found to be significant.

### Latent Class Analysis

Latent class analyses of sexual health-related variables explored several models that ranged from two to five latent classes with model fit statistics shown in Appendix Table III. Based on AIC, BIC and negative log-likelihood as well as considerations of class separation and interpretability, we determined that the four-class model provided the optimal fit for both men and women.

The results in tables 2 and 3 show the conditional probabilities of reporting a behaviour given membership in a certain class for both men and women respectively. With the intent to provide simple and useful characterizations of the latent classes, we termed these four sub-groups using common, gender-neutral names in the UK and their primary descriptors. These four names might aid clinicians in identifying sexual health needs and identifying services in a manner to which each could be most receptive and most relevant. These probabilities formed the basis for the labelling of each class as follows. The first and largest class, made up of 48.9% of the men and 44.9% of the women, was labelled *“Content Caseys”* based on the low likelihoods of reporting higher-risk sexual behaviours, multiple partners and dissatisfaction/distress. Class 2, a smaller class for men (30.9%) than women (44.4%), was labelled “*Infrequent Indigos”* based on the high likelihood that members of this class had not engaged in sex in the last 4 weeks. Class 3, made up of 11.6% of the men and 8.4% of the women, was labelled *“Low-Functioning Lees”* based on the greater probability that members of this class reported a low sexual function score as well as distress regarding their sex life. The fourth class, made up of more men (8.6%) than women (2.3%), was labelled “*Multiple Partner Morgans”* as a result of their high likelihood of having two or more sexual partners in the past year.

**Table 2.** Latent Class Analysis results for Men age 45 and above, N = 1887 (Created by the authors)

| Class Label | Class 1 | Class 2 | Class 3 | Class 4 |
| --- | --- | --- | --- | --- |
|  | Content Caseys | Infrequent | Low-functioning | Multiple-Partner |
|  | (%) | Indigos (%) | Lees (%) | Morgans (%) |
|  | (n = 913) | (n = 583) | (n = 218) | (n = 173) |
| *Overall class size* | *48.86* | *30.94* | *11.65* | *8.62* |
| No sex in last 4 weeks * | 16.63 | **99.39** | 42.48 | 25.51 |
| Had unsafe sex in past year * | 0.01 | 0.01 | 0.12 | **68.60** |
| 2 or more sex partners in past year * | 1.24 | 0.02 | 5.93 | **99.89** |
| Diagnosed with any STI in the last 5 years | 0.78 | 0.02 | 2.34 | 4.30 |
| Paid for sex in past year * | 0.14 | 0.51 | 1.71 | 6.43 |
| Low Sexual Function Score | 4.55 | 5.63 | **98.81** | 17.17 |
| Dissatisfied with sex life | 4.81 | 32.16 | **53.78** | 9.34 |
| Distressed or worried about sex life | 1.13 | 12.19 | **57.40** | 7.42 |

Probabilities greater than 50% are bolded to indicate items that members of a given class were more likely to report. \*Same and opposite sex.

**Table 3.** Latent class analysis results for women age 45 and above, N = 2653 (Created by the authors)

| Class Label | Class 1 | Class 2 | Class 3 | Class 4 |
| --- | --- | --- | --- | --- |
|  | Content Caseys | Infrequent | Low-functioning | Multiple-Partner |
|  | (%) | Indigos (%) | Lees (%) | Morgans (%) |
|  | (n = 1245) | (n = 1144) | (n = 206) | (n = 58) |
| *Overall class size* | *44.91* | *44.38* | *8.41* | *2.30* |
| No sex in last 4 weeks * | 23.02 | **99.98** | 45.72 | 41.28 |
| Had unsafe sex in past year * | 0.01 | 0.01 | 0.04 | 1.74 |
| 2 or more sex partners in past year * | 0.91 | 0.01 | 1.65 | **99.76** |
| Diagnosed with any STI in the last 5 years | 0.30 | 0.65 | 0.18 | 6.27 |
| Paid for sex in past year * | 0.01 | 0.10 | 0.50 | 0.20 |
| Low Sexual Function Score | 6.67 | 6.36 | **96.17** | 37.13 |
| Dissatisfied with sex life | 0.61 | 20.73 | **66.81** | 30.55 |
| Distressed or worried about sex life | 2.69 | 6.90 | **55.49** | 11.46 |

Probabilities greater than 50% are bolded to indicate items that members of a given class were more likely to report. \*Same and opposite sex.

### Demographic, Health, and Lifestyle Characteristics by Classes

The results in Table 4 show the conditional probabilities of reporting a given socio-demographic, general health or help-seeking characteristic given membership in a certain latent class. Due to the similar trends observed among men and women in the creation of these classes, we combined both sexes for this analysis to aid in readability and simplicity. Separate analyses are reported in Appendix Tables IV and V. Broadly, the *Content Caseys* and *Low-Functioning Lees* were very similar in profile, except the *Content Caseys* generally reported better health and less disability. *Infrequent Indigos* were older and more likely to have a disability or a negative view of their own health. *Multiple Partner Morgans* had a better view of their health and reported less disability.

**Table 4.**
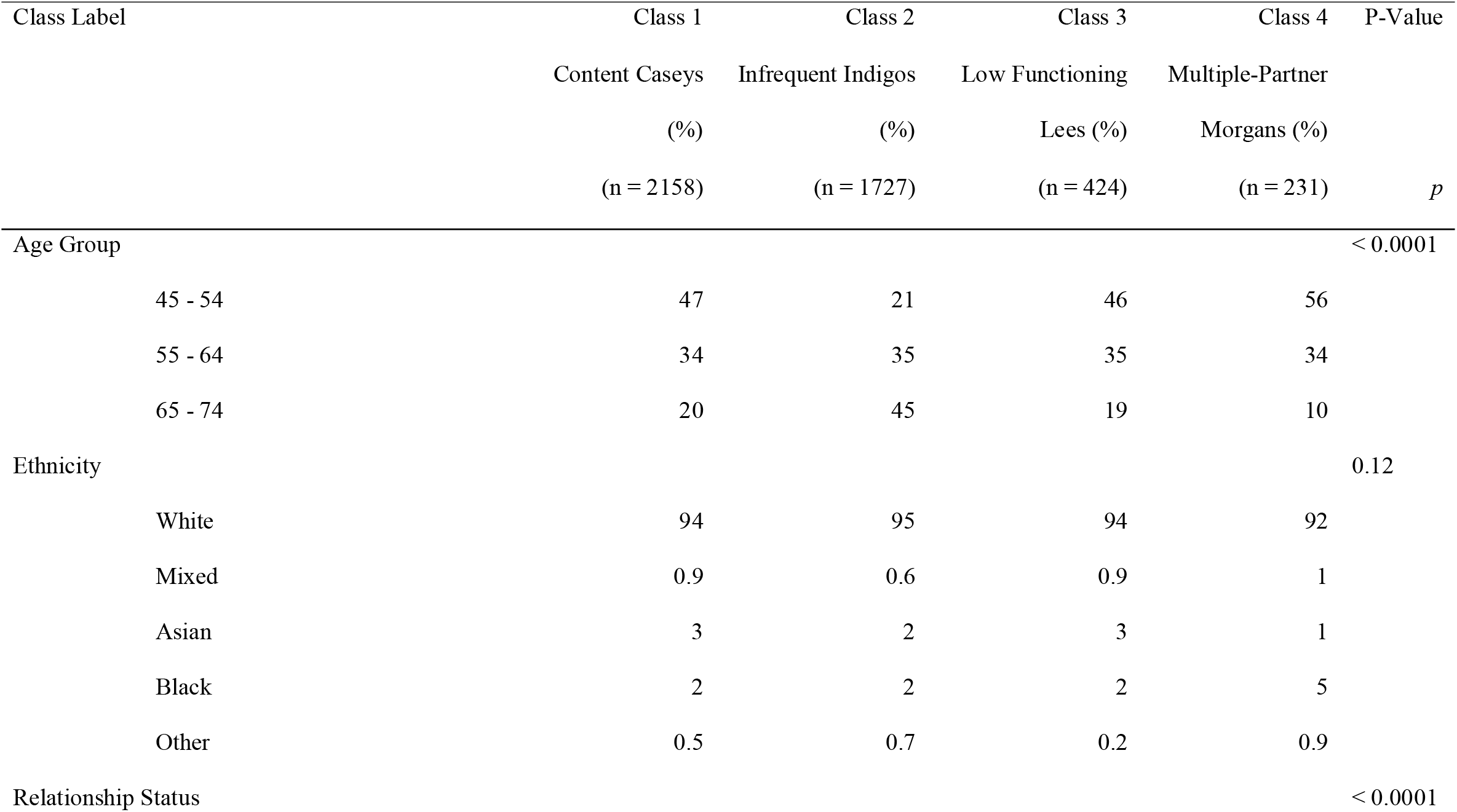
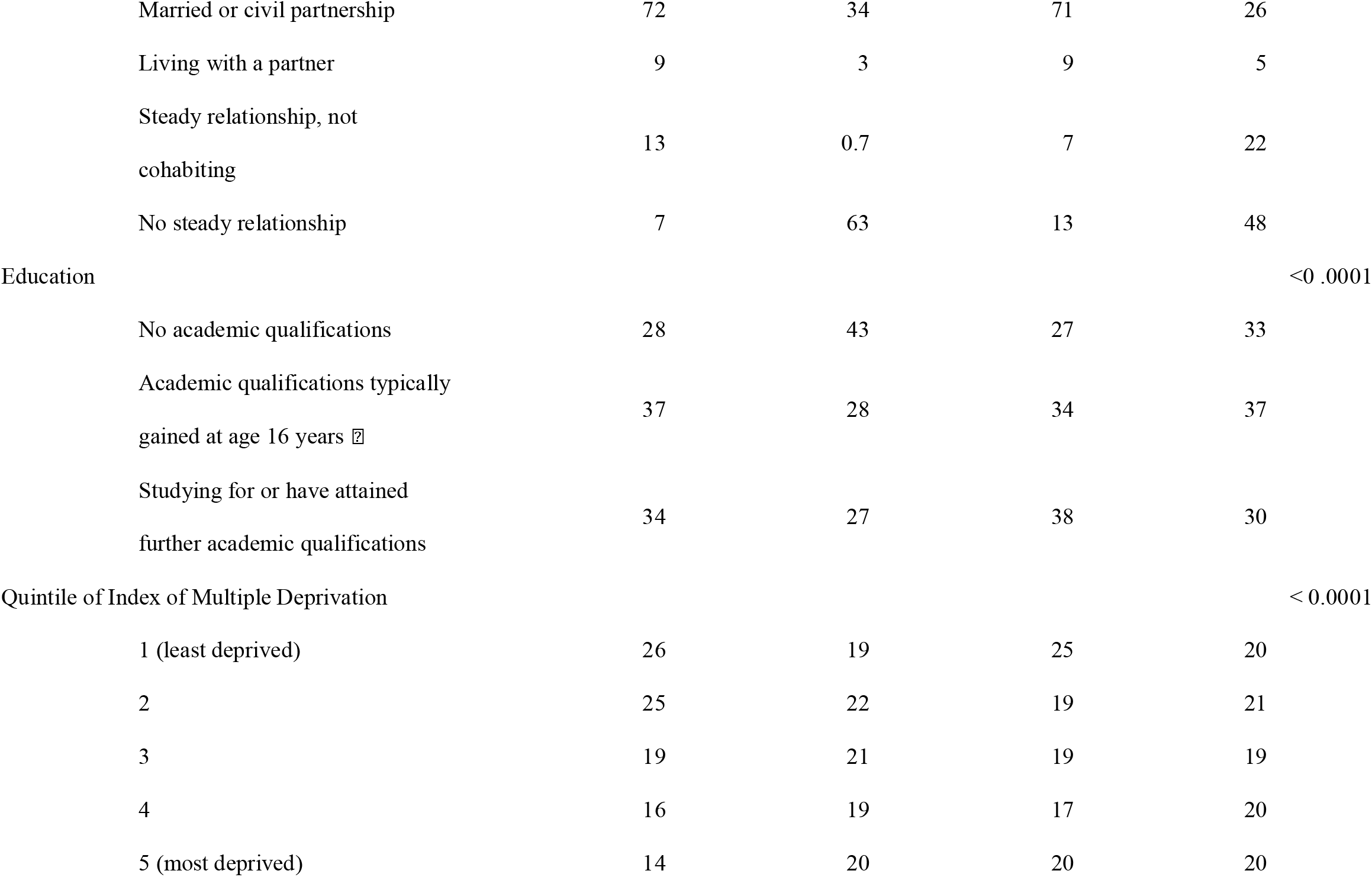
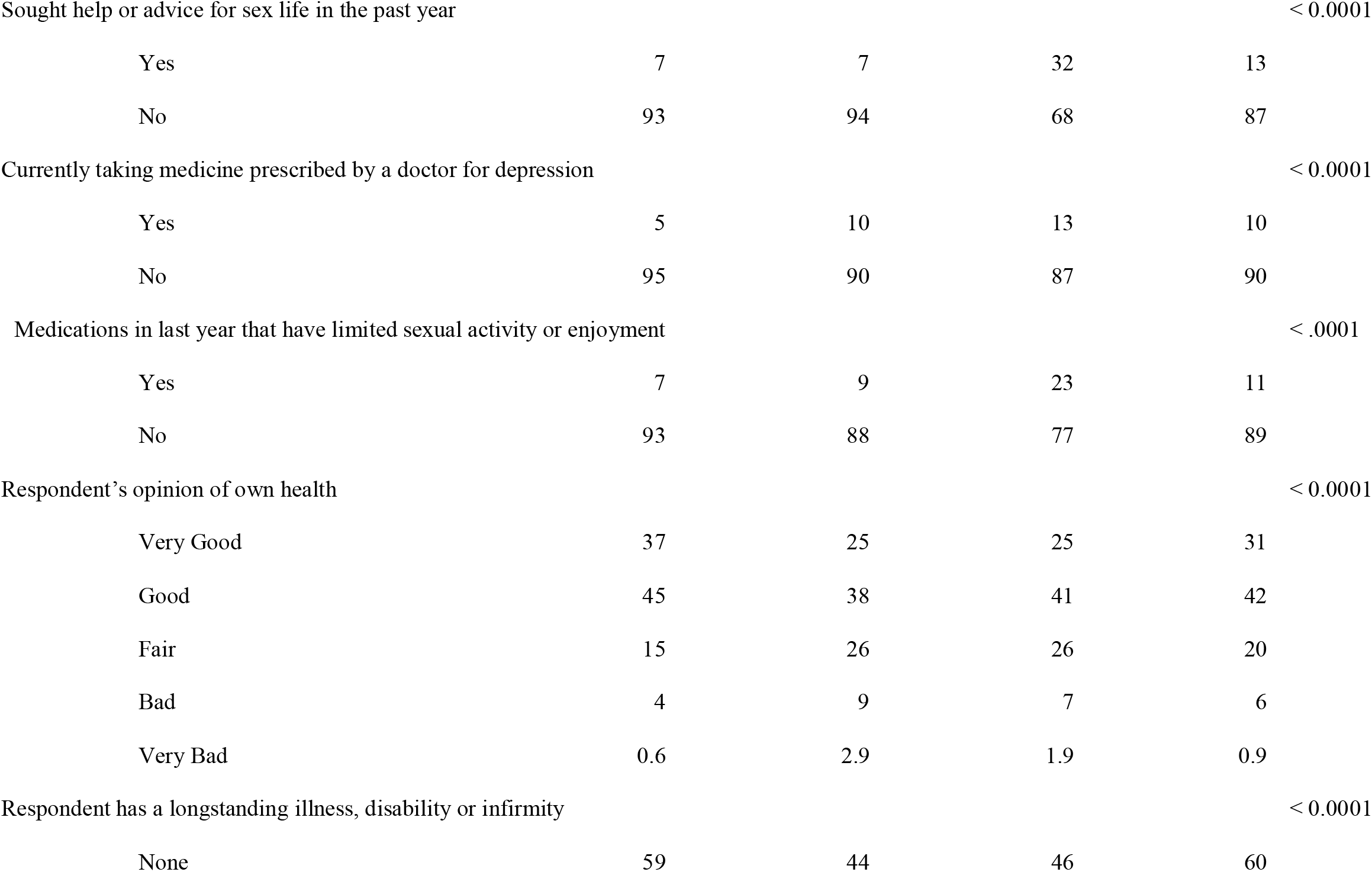
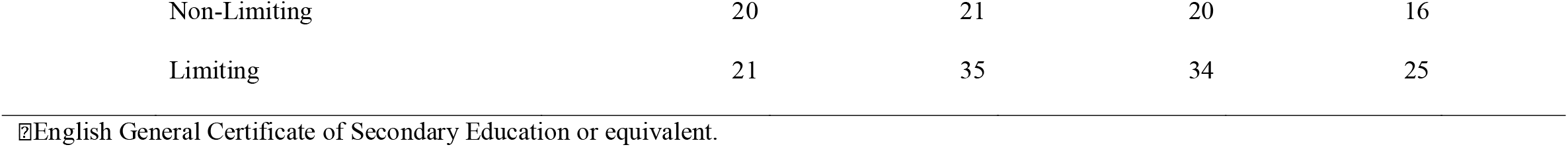
Socio-Demographic, Health and Lifestyle Characteristics by Latent Classes of NATSAL-3 Participants Age 45 and Above, N = 4540 (Created by the authors)

Within the *Infrequent Indigos* group, most were age 65 or above, while among the *Multiple-Partner Morgans* class, the majority were relatively younger (majority 45-54 years). In the *Low-Functioning Lees* class, the majority were married or cohabitating with a partner (71% and 9% respectively) while, conversely, the majority of the *Infrequent Indigos* were in no steady relationship (63%). The *Multiple Partner Morgans* class contained a higher percentage of those in no relationship (48%) than those who were married (26%), and, of the four classes, the largest share of those who were in a relationship without cohabitating (22%). Help-seeking for one’s sexual health was more common for the *Low-Functioning Lees* (32%) and the *Multiple Partner Morgans* (13%) than those in the other two classes. Regarding disability, the *Infrequent Indigos* and *Low-Functioning Lees* classes had a higher proportion of individuals reporting limiting disabilities (35% and 34%) than the *Content Caseys* and *Multiple Partner Morgans* classes (21% and 25%).

## DISCUSSION

Population representative studies that focus on sexual practices, lifestyles, and outcomes among middle-aged and older adults are rare. Our analyses extend the literature by examining sexual lifestyle and outcomes and identifying latent classes among middle-aged and older adults in Britain. These classes were *Content Caseys, Infrequent Indigos, Low-Functioning Lees*, and *Multiple-Partner Morgans*. Overall, about half of all middle-aged and older adults were *Content Caseys* who had good sexual health and reported few issues concerning their sex lives. The other three classes suggested unique needs for sexual health services. A large proportion of these were *Infrequent Indigos* who reported no recent sexual activity, the vast majority of whom are single. Smaller proportions of survey respondents reported issues with sexual function (*Low-Functioning Lees*) and/or multiple partners (*Multiple Partner Morgans*). Interestingly, the lack of differences observed between men and women when creating these classes indicate a strong similarity in the predictive variables measured. Though women were less likely to be assigned to the *Low-Functioning Lees* and *Multiple-Partner Morgans* classes than men, the consistency of the trends among the classes and their respective demographics suggest that the sexual health needs of middle-aged and older men and women are not entirely divergent. While sex should be taken into consideration in middle-aged and older adult care, factors such as relationship status, disability, and behaviour are likely more important in determining the type of need or risk at hand.

The *Low-Functioning Lees* reported the highest levels of distress or worry about their sex lives as well as higher levels of dissatisfaction and relatively high need for professional advice, compared to other classes. Because *Low-Functioning Lees* were associated with more distress, we speculate that these tensions can be compounded among those cohabiting with a partner. This is unlike the *Infrequent Indigos*, whom were cohabitated much less and largely did not report much distress, and *the Content Caseys*, whom had a similar relationship profile but reported less distress and, by definition, higher sexual function scores. In addition to helping improve sexual function, interventions to reduce this psychological stress for *Low-Functioning Lees*, particularly those in long-term relationships, will be essential in taking a comprehensive approach of sexual health for this class. One study analysing high-need patients at a federally qualified health centre in the United States found that their LCA model helped the centre recommend new patients to existing health services.^24^ Using a similar approach, referrals to psychological services, relationship counselling and even sex therapy could improve outcomes for this class.^25^ Additionally, this analysis supports the view that these interventions should include both partners. ^4^

As indicated by the *Low-Functioning Lees* and *Infrequent Indigos* class descriptions, we noticed the concurrence of reporting a disability that limited one’s activities and the reporting of sexual function issues and/or infrequent sex. These classes also observed issues with dissatisfaction and distress and more from these groups tend to have poorer general health and more disability, compared to other classes. These findings on disability and dysfunction are consistent with previous studies which reported on the interrelated nature of general and mental health and sexual wellbeing.^18 26^ Due to these higher levels of distress and dissatisfaction, there is likely a missed opportunity to tailor sexual health services for these two classes. With *Infrequent Indigos* and *Low-Functioning Lees*, our findings suggest that sexual health and wellbeing should be managed alongside general health and that doctors should proactively assess sexual-functioning and sexual lifestyle concerns within the management of disability care. As with LCA-guided referrals, our model can help identify need and raise notice to sexual health issues in clinical sessions that may otherwise be missed.^27^

Individuals in the *Multiple Partner Morgan* class were more likely to have had unsafe sex, to have purchased sex in the past year, and to report a recent STI diagnosis. Combined with their relatively low rates of help-seeking, these behaviours and health history suggest an elevated STI burden among this class and the potential usefulness for STI education services. Literature has alluded to these gaps in education, citing that many older adults came of age prior to the HIV crisis and the proliferation of modern sexual health education campaigns. Therefore, due to age, many in this age range did not receive these services as adults nor as adolescents. ^27-29^ (Pilcher 2005, Terrence Higgins Trust 2018, Choi, Israel et al. 2020) ^27-29^ [27-29] [26-28] [25-27] *Multiple Partner Morgans* therefore could benefit from provider-initiated conversations about safer sex, STI prevention and STI testing during routine clinical visits. As *Multiple Partner Morgans* represent the class of highest risk and could require greater need for diagnostic tests, treatments, and provider consultations, clinics may also find it useful to engage in capacity planning for this class. Such has been done with LCA-enabled growth projections and enhanced screening and triage practices to identify and streamline these patients.^24^

Beyond the clinic, these four classes can be used to gain detailed insight into public health campaigns. LCAs can be used to tailor mass education campaigns, community engagement initiatives or clinical interventions among the different classes.^30^ As this LCA contributes to the research by identifying sets of sexual health needs and areas for more personalised intervention, the potential for the use of these new subgroups for analysis, both in small-scale clinical tailoring and broader intervention research, should be further explored.

## LIMITATIONS

Our use of Natsal-3 survey data does yield to some limitations, most notably the age of the data, as interviews were conducted a decade ago (2010-2012). While these data are now old, they are not necessarily outdated as the relative lack of attention to older adults in sexual health research and the ageing of the British population suggests the relevancy of our analysis today. Though some items, such as online dating, have likely changed since 2012, ELSA noted in 2016 comparable levels of sexual activity and dissatisfaction to Natsal-3,^4^ suggesting minimal changes in sexual behaviour of older adults in recent years.

The original survey did not collect information on intersexed individuals, transgender individuals, nor individuals identifying as a non-binary gender. Though gender identity is now recognized as a meaningful part of sexual wellbeing and our data likely includes these individuals, we report no findings specific to these demographics. Nor does this data represent those individuals in institutionalized living, such as care homes, in which predisposing demographics and frequent lack of sexual health policies ^24^ could mean elevated risk for dysfunction or STIs. In addition, the Natsal-3 study did not survey respondents over 74 years of age, thus rendering a clear, yet incomplete picture of older adults, who can remain sexually active well into their 80s and beyond.^4^

## CONCLUSIONS

Overall, older adults have sex late into their lives and, while many maintain good sexual health, about half experience issues with either low sexual function, distress about sex or an elevated risk for STIs. These reports include those with disabilities. Identifying specific sets of needs and potential areas of risk through our class models can aid researchers and clinicians in tailoring and improving sexual health services to various subgroups of an aging population.

## Supporting information

STROBE Research Checklist

## Data Availability

Entire Natsal-3 data set is available online at https://www.natsal.ac.uk/resources/accessing-data and in the UK Data Service (SN 7799).

http://doi.org/10.5255/UKDA-SN-7799-2

## AUTHOR CONTRIBUTIONS

DW, JDT, HZ, CT, TS, HK, JO, SWP contributed to conceiving the idea. JK and EG co-led the data analysis, interpretation of results and drafted manuscript preparation under the supervision of DW. All authors reviewed the results and provided constructive comments that substantially improved the manuscript. All authors have seen and approved the final version of this manuscript.

## CONFLICTS OF INTEREST

We declare no conflicts of interest.

## FUNDING SUPPORT

This work was jointly supported by Economic and Social Research Council, UK Research and Innovation (UKRI) [grant number: ES/T014547/1], National Natural Science Foundation of China [72061137001], the Natural Science Foundation of China Excellent Young Scientists Fund [82022064], Natural Science Foundation of China Young Scientist Fund [81703278], Special Support Plan for High-Level Talents of Guangdong Province[2019TQ05Y230].

## Notes

### Competing Interest Statement

The authors have declared no competing interest.

### Author Declarations

This study uses publicly available human data collected from 2010-2012 by the Natsal-3 survey study. Data can be found at https://www.natsal.ac.uk/resources/accessing-data and in the UK Data Service (SN 7799). The Natsal-3 study was approved by the Oxfordshire Research Ethics Committee A (reference: 09/H0604/27).

