## Supplemental Tables for "Sexual Behaviours and Sexual Health Among Middle-Aged and Older Adults in Britain: A Latent Class Analysis of The Natsal-3"

**Appendix Table I.** Definitions of key variables used in the analysis and how they were measured

| Variable | Definition of Variable | How variable was asked in Natsal-3 |
| --- | --- | --- |
| Relationship Status | Married or civil partnership if their response was 1 or 2  Living with partner if their response was 3  In steady relationship, not cohabitating if their response was 4  No steady relationship if their response was 5 | At present are you...  1. Married and living with your (husband/wife)  2. In a registered same-sex civil partnership and living with your partner  3. Living with a partner, as a couple (not married or in a civil partnership)  4. In a steady relationship, but not living together  5. None of the above |
| Ethnicity | White if their response was A: 1, 2, or 3 Mixed if their responses was B: 4, 5, 6, or 7 Asian if their response was C: 8, 9, 10, or 11 Black if their response was D: 12, 13, or 14 Chinese if their response was E: 15  Other if their response was E:16 | Participants were asked about which ethnic group they belong to. They could choose: **A. White** 1. British 2. Irish 3. Any Other White background  **B. Mixed** 4. White and Black Caribbean 5. White and Black African 6. White and Asian 7. Any Other Mixed background **C. Asian or Asian British** 8. Indian 9. Pakistani 10. Bangladeshi 11. Any Other Asian background  **D. Black or British Black** 12. Caribbean 13. African 14. Any Other Black background  **E. Chinese or other ethnic group**  15. Chinese 16. Any Other |
| Sexual Identity | Participants were asked to choose from a list sexual orientations on a card and indicate the one that best fits how they think of themselves. | Participants were asked to choose how they best identify themselves. Their options were: 1. Heterosexual / Straight 2. Gay/ Lesbian 3. Bisexual 4. Other |
| Academic Qualifications | No Academic Qualifications if their response was 2.  Academic Qualifications typically achieved at age 16 if gave one of qualifications 5-15 and were no longer in full-time education  Studying for or have attained further academic qualifications if gave one of qualifications 5-15 and were still in full-time education or if gave one of qualifications 1-4.  Those with foreign qualifications are coded as not answered as we do not have information on what level these qualifications were. | Participants were asked: At what age did you complete your continuous full-time education? If you had a 'gap' year between school and university or college please include it as continuous.  Have you passed any exams or got any of the qualifications?  1. Yes 2. No, none  IF Yes THEN Please read down the list and tell me the highest qualification that you have, that is, the first one you come to. INTERVIEWER: Code one only. 1. Degree level qualification 2. A-levels 3. AS level 4. SLC Higher Grade, etc 5. O-level, 1975 or earlier 6. O-level, after 1975 A-C 7. O-level, after 1975 D-E 8. GCSE grades A*-C 9. GCSE grades D-G 10. CSE grade 1, etc 11. CSE grades 2-5, etc 12. CSE Ungraded 13. SLC Lower 14. SUPE Lower or Ordinary 15. School Certificate 16. Foreign qualification |
| Socio-Economic Class | Following the Natsal-3 groupings, participants responses were classified as:  1: Managerial & professional occupations 2: Intermediate occupations 3: Semi-routine / routine occupations 4: Never worked / no job of 10+ hrs/week / not in the last 10 years 5: Student in full-time education | Questions asked here were informed by the International Standard occupation codes. For more information, see the International Standard Occupation Codes (SOC): http://www.ons.gov.uk/ons/guide-method/classifications/archived- standard- classifications/standard-occupational-classification-2000/index.html |
| Limiting Disability | Participants were then grouped in the following way: 1: None 2: Non-limiting 3: Limiting | Participants were asked: "Do you have any long-standing illness, disability or infirmity? By long-standing I mean anything that has troubled you over a period of time, or that is likely to affect you over a period of time?" 1. "Yes" 2. "No"  IF Yes THEN "Does this limit your activities in any way?"  1. "Yes" 2. "No" |
| Opinion of Own Health | Participants were grouped accordingly. | Respondents were asked: 'How is your health in general?' 1: Very good 2: Good 3: Fair 4: Bad 5: Very bad |
| Depression | Respondents could select and record as many conditions as needed. If depression (3) was indicated as one the those treated in the past year then the respondent was classified having 'mentioned' the condition. | Respondents were asked: 'In the last 12 months, that is since (date 12 months ago), have you received treatment from a health professional for any of the medical conditions listed on this card?' 1. Yes  2. No  IF Yes THEN  'Which ones in the last year?' 1. Back ache lasting for 3 months or longer 2. Any other muscle or bone disease lasting for 3 months or longer 3. Depression 4. Any other mental health condition 5. Any other neurological condition, apart from Parkinson’s disease and epilepsy 6 Cancer 7. Any thyroid condition 8. Any (ovarian/testicular) or pituitary condition 9. [WOMEN ONLY] Polycystic ovaries 10. [WOMEN ONLY] I have received IVF or other fertility treatment |
| Quintile of Index of Multiple Deprivation | Respondents were classified: 1 (Least Deprived)  2 3 4 5 (Most Deprived) | Postcodes were used to obtain IMD scores. The adjusted IMD score was generated using a method by Payne and Abel. The individual country IMD scores combined with the co-efficients and residual values from a linear regression of income and employment on the overall IMD score for each country. The combined scores were generated using the most up-to-date scores for each country at the time. These were IMD 2010 for England, IMD 2011 for Wales, and IMD 2009 for Scotland. |
| Sexual Function | Participants were classified, though this scale, as either low functioning, normal functioning, or not sexually active. The lowest quintile class for each gender determined the 'low functioning' measure. | Participants responded to the 17-item Natsal-SF scale, which combines measures of issues related to physical function, mental distress, and satisfaction. While some were asked all 17 items, those whom have never been in a relationship or have not been sexually active in the past year answered only those which applied and modeling techniques were used to estimate the missing answers.   For more information, see Mitchel et al 2011 and Mitchel et al 2012. |
| Medications in Last Year That Have Limited Sexual Activity or Enjoyment | Participants were grouped accordingly. | Participants were asked:  Have you taken any medications in the last year that you feel have limited your sexual or enjoyment in any way? 1 Yes 2 No |
| Dissatisfied with Sex Life | Participants were classified as 'dissatisfied' with their sex life if their response was 4 or 5. | Participants were asked:  I feel satisfied with my sex life  1. Agree strongly  2. Agree  3. Neither agree nor disagree  4. Disagree  5. Disagree strongly |
| Distressed/Worried about Sex Life | Participants were classified as 'distressed' or 'worried" about their sex life if their response was 1 or 2. | Participants were asked:  I feel distressed or worried about my sex life  1. Agree strongly  2. Agree  3. Neither agree nor disagree  4. Disagree  5. Disagree strongly |
| Avoided Sex Because of Own or Partner’s Sexual Difficulties | Participants were classified as avoiding sex if their response was 1 or 2. | Participants were asked:  I have avoided sex because of sexual difficulties, either my own or those of my partner.  1. Agree strongly  2. Agree  3. Neither agree nor disagree  4. Disagree  5. Disagree strongly |
| Happy in Relationship | Participants were classified as "happy in relationship" if their response was 1 or 2. | Participants were asked:  On a scale of 1 to 7, where 1 means very happy and 7 means very unhappy, how happy or unhappy are you with your relationship with your partner, all things considered? 1..7 |
| STI diagnosis | Respondents who reported an STI diagnosis and specified 1 or 2 for the follow up questions, were recorded as having an STI diagnosis in the past 5 years. | Participants were asked: Have you ever been told by a doctor or other healthcare professional that you had any of the following?  Please select any that you have had, even if not transmitted by sex (women only: Thrush). If more than one, press the space bar between each number. If you have not had any, please select 'none of these' at the bottom of the list.  1. Chlamydia 2. Gonorrhoea 3. Genital warts (venereal warts) 4. Syphilis 5. Trichomonas vaginalis (Trich, TV) 6. Herpes (genital herpes) 7. Pubic lice / crabs 8. Hepatitis B 9. (Men only:) NSU (Non Specific Urethritis), NGU (Non Gonococcal Urethritis) 10. (Men only:) Epididymitis 11. (Women only:) Pelvic Inflammatory Disease (PID, salpingitis) 12. (Women only:) Vaginal thrush (Candida, Yeast infection) 13. (Women only:) Bacterial vaginosis 14. Yes, but can't remember which 15. None of these  Respondents that selected 1-9, or reported a diagnosis of HIV, were then asked to specify "when were you last told by a doctor or healthcare professional that you had ___?"  1. Less than 1 year ago  2. Between 1 and 5 years ago  3. Between 5 and 10 years ago  4. More than 10 years ago |
| For more information on definitions, phrasings, and other variables consult the Natsal-3 questionnaire and codebook, available at www.natsal.ac.uk | | |

**Appendix Table II. Table 2.** Sexual partners, practices, behaviours and opinions of male and female participants in NATSAL-3, by age group, 2012, UK, N = 15162

| Class Label | | Men (%) | | | | Women (%) | | | |
| --- | --- | --- | --- | --- | --- | --- | --- | --- | --- |
|  |  | < 45 | 45-54 | 55 - 64 | 65 - 74 | < 45 | 45-54 | 55 - 64 | 65 - 74 |
|  |  | (n = 4060) | (n = 794) | (n = 772) | (n = 667) | (n = 5842) | (n = 1123) | (n = 1030) | (n = 874) |
| Number of partners in last year | |  |  |  |  |  |  |  |  |
|  | 0-1 | 72 | 82 | 82 | 86 | 78 | 89 | 93 | 91 |
|  | 2+ | 26 | 14 | 10 | 4 | 19 | 6 | 2 | 1 |
| Partners | |  |  |  |  |  |  |  |  |
|  | Had a concurrent partnership in the last 5 years | 18 | 14 | 9 | 4 | 13 | 6 | 2 | 0.6 |
|  | Has at least one new partner in past year | 35 | 16 | 13 | 5 | 27 | 12 | 5 | 2 |
|  | Used internet to find sexual partner in last year * | 6 | 4 | 3 | 2 | 3 | 2 | 2 | 1 |
| Number of occasions of sex § in last 4 weeks | |  |  |  |  |  |  |  |  |
|  | 0 | 30 | 29 | 42 | 59 | 27 | 36 | 59 | 75 |
|  | 1-2 | 18 | 22 | 20 | 16 | 18 | 21 | 14 | 11 |
|  | 3-4 | 15 | 17 | 12 | 7 | 16 | 15 | 11 | 3 |
|  | 5+ | 32 | 25 | 14 | 7 | 30 | 18 | 9 | 2 |
| Sexual Practices | |  |  |  |  |  |  |  |  |
|  | Vaginal sex in last year | 84 | 83 | 70 | 52 | 84 | 79 | 57 | 34 |
|  | Given or recieved oral sex in past year | 76 | 69 | 50 | 28 | 74 | 60 | 34 | 17 |
|  | Anal sex in past year | 17 | 13 | 7 | 3 | 15 | 7 | 3 | 3 |
|  | Genital contact without intercourse in past year | 72 | 64 | 53 | 34 | 73 | 61 | 39 | 26 |
| Masturbation | |  |  |  |  |  |  |  |  |
|  | Masturbated in last month | 75 | 62 | 49 | 30 | 39 | 36 | 18 | 9 |
| Risky Sexual Behaviours | |  |  |  |  |  |  |  |  |
|  | Had unsafe sex § in past year † | 6 | 7 | 5 | 2 | 6 | 3 | 1 | 0.5 |
|  | Paid for sex § in last year | 1 | 1 | 2 | 0.5 | 0.0 | 0.1 | 0.0 | 0.1 |
| STI Diagnosis | |  |  |  |  |  |  |  |  |
|  | Diagnosed with any STI in the last five years | 6 | 2 | 0.4 | 0.6 | 7 | 1.2 | 0.3 | 0.3 |
| Opinion on Sex Life | |  |  |  |  |  |  |  |  |
|  | Happy in relationship | 38 | 43 | 35 | 28 | 38 | 39 | 29 | 15 |
|  | Dissatisfied with sex life | 16 | 18 | 18 | 17 | 12 | 16 | 15 | 10 |
|  | Worried about sex life | 9 | 10 | 13 | 10 | 11 | 12 | 9 | 7 |
|  | Avoided sex because of own or partner's sexual difficulties | 9 | 12 | 16 | 20 | 10 | 15 | 20 | 16 |
| Sexual Function | |  |  |  |  |  |  |  |  |
|  | Normal function | 71 | 70 | 51 | 40 | 71 | 64 | 45 | 29 |
|  | Low function | 15 | 16 | 20 | 15 | 15 | 19 | 17 | 9 |
|  | Not sexually active | 9 | 11 | 20 | 35 | 9 | 13 | 34 | 52 |

All estimates are weighted. All participants (denominators vary across variables because of item non-response). Vaginal sex is defined as a man’s penis in a woman’s vagina. Oral Sex is defined as mouth on a partner’s genital area. Anal sex is defined as man’s penis in a partner’s anus. *If at least one partner ever. †Unsafe defined as two or more partners and no condom use in the last year. §Heterosexual/same-sex vaginal, oral or anal sex. ‡Excluding Thrush

Appendix Table III: Latent class analyses Model Fit Statistics for Participants Aged 45 and above, N = 4540

|  | Number of Classes | AIC | BIC | Negative Log-Likelihood |
| --- | --- | --- | --- | --- |
| Men | 2 | 12,835.0 | 12,973.6 | 6,392.52 |
|  | 3 | 12,249.1 | 12,459.7 | 6,086.53 |
|  | **4** | **11,803.6** | **12,086.3** | **5,850.81** |
|  | 5 | 11,766.2 | 12,121.0 | 5,819.08 |
| Women | 2 | 14,840.3 | 14,987.4 | 7,395.17 |
|  | 3 | 14,139.8 | 14,363.4 | 7,031.89 |
|  | **4** | **13,842.1** | **14,142.2** | **6,870.04** |
|  | 5 | 13,804.3 | 14,180.9 | 6,838.16 |

Figures in bold show that the fit for the four-class model was selected for both men and women. AIC Alkaline Information Criterion, BIC Bayesian Information Criterion, -LogLikelihood Negative log-likelihood

**Appendix Table IV.** Socio-demographic, health and lifestyle characteristics of male participants, N = 1887

| Class Label | | Class 1 | Class 2 | Class 3 | Class 4 | P-Value |
| --- | --- | --- | --- | --- | --- | --- |
|  |  | Content Caseys (%) | Occasional Ollies (%) | Low Function Frankies (%) | Multiple-Partner Morgans (%) |  |
|  |  | (n = 913) | (n = 583) | (n = 218) | (n = 173) |  |
| Age Group | |  |  |  |  | < .0001 |
|  | 45 - 54 | 44 | 23 | 38 | 53 |  |
|  | 55 - 64 | 33 | 32 | 38 | 36 |  |
|  | 65 - 74 | 23 | 46 | 25 | 11 |  |
| Ethnicity | |  |  |  |  | < .0001 |
|  | White | 95 | 96 | 94 | 93 |  |
|  | Mixed | 1 | 1 | 1 | 1 |  |
|  | Asian | 3 | 2 | 3 | 2 |  |
|  | Black | 2 | 0 | 2 | 4 |  |
|  | Other | 0 | 0 | 0 | 1 |  |
| Relationship Status | |  |  |  |  | < .0001 |
|  | Married or civil partnership | 74 | 37 | 73 | 25 |  |
|  | Living with a partner | 10 | 4 | 10 | 5 |  |
|  | Steady relationship, not cohabiting | 12 | 1 | 6 | 23 |  |
|  | No steady relationship | 5 | 58 | 11 | 46 |  |
| Education | |  |  |  |  | < .0001 |
|  | No academic qualifications | 27 | 46 | 29 | 34 |  |
|  | Academic qualifications typically gained at age 16 years ☨ | 35 | 22 | 31 | 34 |  |
|  | Studying for or have attained further academic qualifications | 37 | 31 | 39 | 32 |  |
| Quintile of Index of Multiple Deprivation | |  |  |  |  | < .0001 |
|  | 1 (least deprived) | 25 | 18 | 26 | 19 |  |
|  | 2 | 25 | 22 | 19 | 21 |  |
|  | 3 | 20 | 20 | 20 | 19 |  |
|  | 4 | 16 | 20 | 15 | 19 |  |
|  | 5 (most deprived) | 14 | 19 | 19 | 22 |  |
| Sought help or advice for sex life in the past year | |  |  |  |  | < .0001 |
|  | Yes | 8 | 10 | 34 | 13 |  |
|  | No | 92 | 90 | 66 | 87 |  |
| Currently taking medicine prescribed by a doctor for depression | | |  |  |  | < .0001 |
|  | Yes | 3 | 8 | 10 | 7 |  |
|  | No | 97 | 92 | 90 | 93 |  |
| Medications in last year that have limited sexual activity or enjoyment | | |  |  |  | < .0001 |
|  | Yes | 9 | 18 | 28 | 11 |  |
|  | No | 91 | 79 | 71 | 89 |  |
| Respondent’s opinion of own health | |  |  |  |  | < .0001 |
|  | Very Good | 38 | 19 | 21 | 33 |  |
|  | Good | 44 | 39 | 42 | 41 |  |
|  | Fair | 14 | 29 | 29 | 20 |  |
|  | Bad | 3 | 10 | 6 | 6 |  |
|  | Very Bad | 0 | 3 | 2 | 1 |  |
| Respondent has a longstanding illness, disability or infirmity | | |  |  |  | < .0001 |
|  | None | 60 | 40 | 44 | 62 |  |
|  | Non-Limiting | 22 | 22 | 21 | 15 |  |
|  | Limiting | 19 | 37 | 35 | 23 |  |

Probabilities greater than 50% are bolded to indicate items that members of a given class were more likely to endorse. ☨English General Certificate of Secondary Education or equivalent.

**Appendix Table V.** Socio-demographic, health and lifestyle characteristics of female participants, N = 2653

| Class Label | | Class 1 | Class 2 | Class 3 | Class 4 | P-Value |
| --- | --- | --- | --- | --- | --- | --- |
|  |  | Content Caseys (%) | Infrequent Indigos (%) | Low Functioning Lees (%) | Multiple-Partner Morgans (%) |  |
|  |  | (n = 2158) | (n = 1727) | (n = 424) | (n = 231) | *p* |
| Age Group | |  |  |  |  |  |
|  | 45 - 54 | 49 | 19 | 54 | 64 |  |
|  | 55 - 64 | 34 | 37 | 33 | 29 |  |
|  | 65 - 74 | 17 | 44 | 13 | 7 | < 0.0001 |
| Ethnicity | |  |  |  |  |  |
|  | White | 94 | 95 | 94 | 92 |  |
|  | Mixed | 0.9 | 0.6 | 0.9 | 1 |  |
|  | Asian | 3 | 2 | 3 | 1 |  |
|  | Black | 2 | 2 | 2 | 5 |  |
|  | Other | 0.5 | 0.7 | 0.2 | 0.9 | 0.12 |
| Relationship Status | |  |  |  |  |  |
|  | Married or civil partnership | 72 | 34 | 71 | 26 |  |
|  | Living with a partner | 9 | 3 | 9 | 5 |  |
|  | Steady relationship, not cohabiting | 13 | 0.7 | 7 | 22 |  |
|  | No steady relationship | 7 | 63 | 13 | 48 | < 0.0001 |
| Education | |  |  |  |  |  |
|  | No academic qualifications | 28 | 43 | 27 | 33 |  |
|  | Academic qualifications typically gained at age 16 years ☨ | 37 | 28 | 34 | 37 |  |
|  | Studying for or have attained further academic qualifications | 34 | 27 | 38 | 30 | <0 .0001 |
| Quintile of Index of Multiple Deprivation | |  |  |  |  |  |
|  | 1 (least deprived) | 26 | 19 | 25 | 20 |  |
|  | 2 | 25 | 22 | 19 | 21 |  |
|  | 3 | 19 | 21 | 19 | 19 |  |
|  | 4 | 16 | 19 | 17 | 20 |  |
|  | 5 (most deprived) | 14 | 20 | 20 | 20 | < 0.0001 |
| Sought help or advice for sex life in the past year | |  |  |  |  |  |
|  | Yes | 7 | 7 | 32 | 13 |  |
|  | No | 93 | 94 | 68 | 87 | < 0.0001 |
| Currently taking medicine prescribed by a doctor for depression | | |  |  |  |  |
|  | Yes | 5 | 10 | 13 | 10 |  |
|  | No | 95 | 90 | 87 | 90 | < 0.0001 |
| Medications in last year that have limited sexual activity or enjoyment | | |  |  |  |  |
|  | Yes | 7 | 9 | 23 | 11 |  |
|  | No | 93 | 88 | 77 | 89 | < .0001 |
| Respondent’s opinion of own health | |  |  |  |  |  |
|  | Very Good | 37 | 25 | 25 | 31 |  |
|  | Good | 45 | 38 | 41 | 42 |  |
|  | Fair | 15 | 26 | 26 | 20 |  |
|  | Bad | 4 | 9 | 7 | 6 |  |
|  | Very Bad | 0.6 | 3 | 2 | 0.9 | < 0.0001 |
| Respondent has a longstanding illness, disability or infirmity | | |  |  |  |  |
|  | None | 59 | 44 | 46 | 60 |  |
|  | Non-Limiting | 20 | 21 | 20 | 16 |  |
|  | Limiting | 21 | 35 | 34 | 25 | < 0.0001 |

Probabilities greater than 50% are bolded to indicate items that members of a given class were more likely to endorse. ☨English General Certificate of Secondary Education or equivalent.
